# Efficacy and Safety of a Voltage-Guided Stepwise Strategy for Pulmonary Vein Isolation: A Randomized Comparison With Conventional Circumferential Antral Pulmonary Vein Isolation

**DOI:** 10.64898/2026.01.21.26344569

**Authors:** Dong-Hyeok Kim, Yong-Soo Baek, Do Young Kim, Gyo-Seung Hwang, Dae-In Lee, Kwang-No Lee

**Author notes:** Corresponding author: Dae In Lee, MD, Division of Cardiology, Department of Internal Medicine, Korea University College of Medicine and Korea University Guro Hospital, 148 Gurodong-ro, Guro-gu, Seoul 08308, Republic of Korea, 08308; Kwang-No Lee, MD, PhD, Department of Cardiology, Ajou University School of Medicine and Ajou University Hospital, World cup-ro 164, Yeongtong-gu, Suwon, Gyeonggi-do, Republic of Korea, 16499.

## Abstract

**Background:** Catheter ablation is an established rhythm control therapy for atrial fibrillation. However, as the extent of ablation increases, the risk of complications may also rise. This has motivated strategies that achieve pulmonary vein isolation with less lesion creation while preserving safety and effectiveness.

**Methods:** In this prospective, multicenter, randomized non-inferiority trial, 130 patients undergoing first-time ablation for paroxysmal or non-paroxysmal atrial fibrillation were assigned 1:1 to a voltage-guided stepwise pulmonary vein isolation approach or conventional circumferential antral pulmonary vein isolation with voltage blinded to operators. The primary end point was recurrence of atrial tachyarrhythmia within 12 months after ablation.

**Results:** At 12 months, recurrence occurred in 23/65 (35.4%) in the stepwise group versus 13/65 (20.0%) in the control group (risk difference 15.4 percentage points; 90% confidence interval, 2.7–28.1), and non-inferiority was not demonstrated (one-sided P=0.520). The treatment group had a higher risk of recurrent atrial tachyarrhythmia than the control group (hazard ratio, 2.05; 95% confidence interval, 1.04–4.06), with longer procedure times and more frequent acute pulmonary vein reconnection after the initial lesion set. The treatment group had fewer acute complications than the control group (1.5% versus 9.2%; P=0.115), and esophageal thermal injury was observed only in the control group (3 cases).

**Conclusions:** Voltage-guided stepwise pulmonary vein isolation failed to demonstrate non-inferiority to conventional circumferential antral pulmonary vein isolation for 12-month atrial tachyarrhythmia recurrence.

ClinicalTrials.gov ID: NCT07354737

## Introduction

Catheter ablation is an established rhythm control therapy for atrial fibrillation, and pulmonary vein isolation is the cornerstone lesion set for both paroxysmal and non-paroxysmal disease.^1,2^ However, conventional ablation delivers lesions around the pulmonary vein, including the posterior left atrial region, placing the esophagus at risk for collateral thermal injury.^3^ Strategies that reduce lesion burden are therefore attractive, but they must still achieve and maintain durable electrical pulmonary vein isolation to prevent reconnection and recurrent atrial tachyarrhythmia.^4–7^

Recent advances in multielectrode technology have enabled high-density electroanatomical mapping. Grid-pattern high-density mapping catheters record bipolar signals in two orthogonal directions at each location and annotate the higher amplitude signal, which can reduce wavefront direction dependence compared with other catheters that rely on fixed electrode pair orientations.^8^ As a result, high-density voltage mapping can more reliably define regional electrogram amplitude patterns and identify areas of preserved myocardium that generate near field depolarization signals of higher amplitude, consistent with a stronger local electrical source.^9^ This can support prioritizing ablation at regions with higher bipolar voltage and extending lesions only as needed to achieve isolation. In contrast, prior segmental approaches used voltage information mainly to identify discrete pulmonary vein potentials based on earliest timing or electrogram morphology.^4–7^

Accordingly, we designed this multicenter randomized trial with a non-inferiority framework. We tested whether a voltage-guided stepwise pulmonary vein isolation strategy that prioritizes ablation at regions with higher bipolar voltage can reduce lesion burden, reflected by the amount of atrial tissue ablated, while maintaining arrhythmia outcomes comparable to conventional circumferential antral isolation. We also explored how regional voltage characteristics relate to the voltage level at which isolation is achieved.

## Methods

### Study Design and Population

This prospective, multicenter, randomized controlled trial compared voltage-guided stepwise pulmonary vein isolation (treatment group) with conventional circumferential antral pulmonary vein isolation (control group) in patients with paroxysmal and non-paroxysmal atrial fibrillation (Figure 1). After providing written informed consent, eligible patients were randomized in a 1:1 ratio. The study protocol was approved by the institutional review boards of all participating centers.

**Figure 1.**
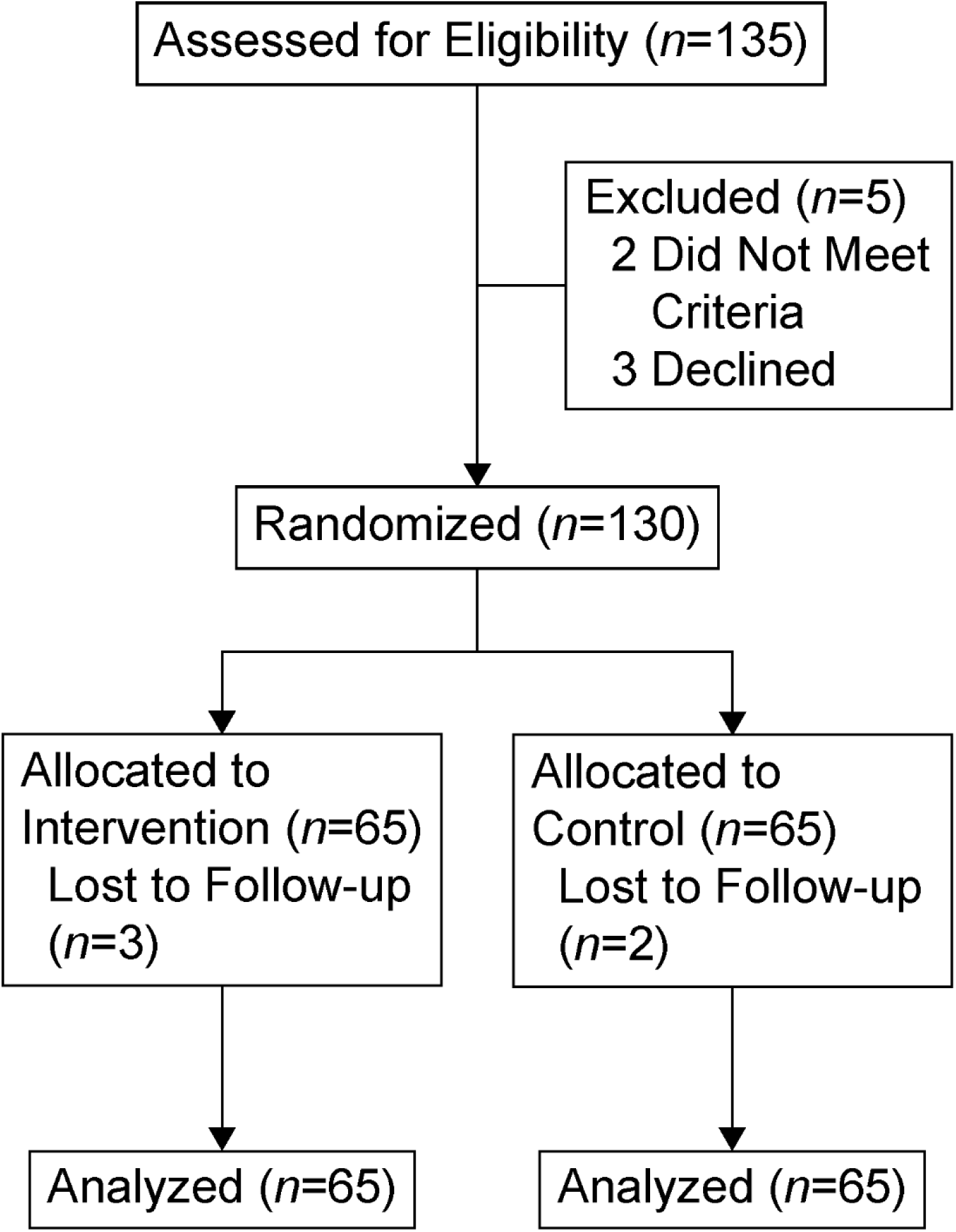
Study Flowchart

Patients were eligible if they were 20 years or older and scheduled for first-time catheter ablation of atrial fibrillation. Key exclusion criteria included a history of prior catheter or surgical ablation for atrial arrhythmia, myocardial infarction within 3 months, stroke within 6 months, New York Heart Association class III or IV heart failure, contraindications to oral anticoagulant therapy, pregnancy, or any medical condition deemed to interfere with procedural safety or study participation.

### Sample Size Calculation

The sample size was calculated based on a non-inferiority design comparing 1-year recurrence rates between the two groups. The recurrence-free rate in the reference group was assumed to be 80 percent. The treatment group was expected to have a similar rate. A non-inferiority margin of 15 percent was applied, using a one-sided significance level of 0.05 and a target power of 70 percent.

### Randomization and Blinding

Randomization was performed at the coordinating center immediately after informed consent using a computer-generated allocation sequence. Allocation was stratified by participating center using permuted blocks of variable sizes to ensure balanced distribution across sites. The allocation sequence was securely managed, and group assignments were concealed until enrollment. Assignment results were disclosed to site investigators only immediately prior to the ablation procedure. Operators were not blinded to group assignment.

### Periprocedural Management and Ablation Procedure

All patients underwent cardiac computed tomography or magnetic resonance imaging to construct a three-dimensional geometry of the left atrium. Oral anticoagulation was maintained for at least three weeks before the procedure. Class I antiarrhythmic drugs were discontinued at least two weeks before, and Class III agents, including amiodarone, at least eight weeks before the procedure.

Procedures were performed using a three-dimensional electroanatomical mapping system (EnSite NavX, Abbott). Geometry of the left atrium was created using a high-density multielectrode mapping catheter (Advisor HD Grid Sensor Enabled Mapping Catheter, Abbott), which recorded both unipolar and bipolar signals. Unipolar voltage was defined as the peak-to-peak amplitude of the unipolar electrogram measured within a 200–300 ms window immediately preceding QRS onset to minimize ventricular far-field influence. Bipolar signals were collected in two orientations by pairing electrodes located along the same spline and across adjacent splines, allowing assessment of local voltage in both parallel and perpendicular directions. For each mapping point, the system selected the higher amplitude from orthogonal bipolar pairs for voltage annotation. This approach was applied to reduce the influence of signal direction on the measured values.

In the treatment group, operators were allowed to view the real-time voltage map throughout the procedure and could modify the lesion set in response to observed voltage characteristics (Central illustration). In the control group, operators performed ablation guided solely by the anatomical geometry, without access to real-time voltage data, although voltage recordings were retained for later analysis.

In the treatment group, ablation was initiated point-by-point at sites with bipolar voltage of 6.0 mV or higher, based on a predefined antral line. Pulmonary veins were treated in a standardized sequence of left superior, left inferior, right superior, and right inferior. For each superior pulmonary vein, carina ablation was performed as needed to achieve isolation. If pulmonary vein isolation was not achieved, the voltage threshold was lowered in 1.0 mV increments to a minimum of 1.0 mV. If isolation could not be achieved even at 1.0 mV, previously ablated points were connected to form a continuous line. Additional potential-guided ablation was performed if necessary. In the control group, a circumferential antral lesion set was delivered around each pair of ipsilateral veins. If isolation was incomplete, potential-guided ablation was also permitted. Energy settings and lesion delivery techniques were identical across both groups.

The procedural end point was defined as complete elimination or dissociation of all pulmonary vein signals, absence of dormant conduction following intravenous adenosine (6–18 mg), and absence of non-pulmonary vein triggers after high-dose isoproterenol infusion (10–20 µg/min).

### Postprocedural Management and Follow-up

Antiarrhythmic drugs were continued for six to twelve months at the discretion of the treating physician. Oral anticoagulation was maintained for at least two months and extended based on each patient’s CHA₂DS₂-VASc score. Follow-up evaluations were conducted at three, six, nine, and twelve months and every six months thereafter, including 12-lead electrocardiograms and 24-or 72-hour Holter monitoring.

### Study End Points

The primary end point was recurrence of any atrial tachyarrhythmia, including atrial fibrillation, atrial flutter, or atrial tachycardia, within twelve months following the index ablation. Recurrence was defined as an episode confirmed on electrocardiography or lasting at least 30 seconds on Holter monitoring. A blanking period of three months was applied, during which arrhythmic episodes were not counted as recurrence.

Time to first documented recurrence was also analyzed using survival methods, including Kaplan-Meier estimates and Cox regression modeling. These time-to-event analyses were performed to assess the distribution of recurrence over the follow-up period and to identify relevant clinical or procedural predictors.

Safety outcomes included acute procedural complications, which were prospectively recorded and comprised death, cardiac tamponade requiring pericardiocentesis or surgical drainage, new permanent pacemaker implantation, phrenic nerve palsy, clinically significant hematoma at the vascular access site, ischemic stroke or systemic embolism, and esophageal thermal injury. To systematically screen for esophageal thermal injury, diagnostic esophagoscopy was performed between the first and third day after the ablation procedure. Esophageal thermal injury was defined as any endoscopically detected abnormality along the esophageal segment adjacent to the posterior left atrial wall that was considered compatible with thermal injury, including mucosal erythema, ulceration involving any esophageal wall layer, or any degree of fistulous communication such as atrio-esophageal fistula. Lesions clearly attributable to other causes, such as reflux, were not classified as esophageal thermal injury.

### Statistical Analysis

Statistical analysis followed the intention-to-treat principle. Continuous variables were compared using the Student t test or the Mann-Whitney U test, depending on data distribution. The Mann–Whitney U test was applied when normality or homoscedasticity assumptions for the t test were not met, or when variables were expressed as medians with interquartile ranges because of skewed distributions. Categorical variables were analyzed using chi-square tests or Fisher’s exact test, as appropriate. The chi-square test was used as the primary method of comparison. Fisher’s exact test was applied when expected cell counts were small. For variables with three or more categories, we additionally used the likelihood ratio chi-square approach.

As an exploratory assessment of segment-level electrogram characteristics, the agreement between bipolar and unipolar voltages was evaluated using Bland–Altman analysis, and Spearman’s correlation coefficients (ρ) were calculated. To compare the bipolar–unipolar relationship between groups, a linear mixed-effects model was additionally applied, specifying subject identifier as a random intercept and including bipolar voltage, group, and their interaction as fixed effects.

In a separate exploratory analysis, we examined the relationship between pre-ablation segmental maximal bipolar voltage and the bipolar voltage threshold at which pulmonary vein isolation was achieved. To illustrate this association, segment-level scatter plots with fitted regression lines were generated.

In an additional exploratory analysis, we sought to identify clinical and anatomical factors associated with variation in the isolation voltage threshold. Because the isolation voltage threshold is influenced both by fixed covariates and substantial between-subject variability, we employed linear mixed-effects modeling to estimate adjusted marginal means for variables of interest while accounting for subject-specific random effects. This approach allowed us to evaluate whether the expected isolation threshold differed across pulmonary veins, atrial fibrillation phenotype, and medication exposure, while recognizing that the magnitude of random intercepts limited the feasibility of deriving reliable patient-level threshold estimates.

To facilitate interpretation and visualization of these adjusted effects, marginal means with 95% confidence intervals were displayed using error-bar plots, and subject-specific random intercepts from the mixed-effects model were plotted to illustrate the extent of interindividual variability. These analyses were intended to descriptively characterize sources of variability in isolation voltage threshold and to contextualize the role of individual patient effects, rather than to provide definitive inferential conclusions.

Atrial tachyarrhythmia-free survival was estimated with the Kaplan-Meier method and compared using the log-rank test. Multivariable Cox proportional hazards models were applied to identify predictors of recurrence and assess subgroup interactions. Statistical significance was defined as a two-sided p value less than 0.05. For the primary non-inferiority evaluation, the 1-year atrial tachyarrhythmia recurrence rate was compared between groups using an absolute risk-difference approach (treatment group minus control group). A non-inferiority margin of 15 percentage points was prespecified. Non-inferiority was concluded if the upper bound of the one-sided 95% confidence interval (equivalent to a two-sided 90% confidence interval) for the risk difference was less than the prespecified margin. The corresponding one-sided non-inferiority p value was calculated using a score-based method for independent proportions. All analyses were performed with SPSS software, version 26.0 (IBM Corp., Armonk, NY, USA).

## Results

### Study population and baseline characteristics

A total of 130 patients were included in the analysis (treatment group, n=65; control group, n=65). Baseline characteristics were well balanced between groups (Table 1). The mean age of the overall cohort was 59.2 ± 9.7 years, 23.8% were women, 53.1% had paroxysmal atrial fibrillation, and the anteroposterior left atrial diameter on transthoracic echocardiography was 42.7 ± 6.2 mm. No clinically meaningful differences were observed between treatment groups in demographic or clinical variables.

**Table 1.** Baseline characteristics.

|  | Treatment (n=65) | Control (n=65) | p-value |
| --- | --- | --- | --- |
| Female, n | 14 (21.5%) | 17 (26.2%) | 0.537 |
| Age, years | 59.5 ± 9.9 | 59.0 ± 9.6 | 0.746 |
| Weight, kg | 74.3 ± 13.4 | 73.8 ± 12.7 | 0.813 |
| Height, cm | 168.1 ± 8.7 | 168.0 ± 8.9 | 0.990 |
| Paroxysmal Atrial Fibrillation, n | 38 (58.5%) | 31 (47.7%) | 0.219 |
| Sick Sinus Syndrome, n | 6 (9.2%) | 6 (9.2%) | 1.000 |
| Heart Failure, n | 22 (33.8%) | 21 (32.3%) | 0.852 |
| Hypertension, n | 32 (49.2%) | 38 (58.5%) | 0.291 |
| Diabetes Mellitus, n | 11 (16.9%) | 14 (21.5%) | 0.504 |
| Prior Ischemic Stroke, n | 3 (4.6%) | 3 (4.6%) | 1.000<sup>*</sup> |
| Prior Intracranial Hemorrhage, n | 0 (0.0%) | 0 (0.0%) | 1.000 |
| Prior Systemic Embolism, n | 0 (0.0%) | 1 (1.5%) | 1.000<sup>*</sup> |
| Coronary Arterial Disease, n | 5 (7.7%) | 3 (4.6%) | 0.718<sup>*</sup> |
| CHA<sub>2</sub>DS<sub>2</sub>-VASc Score | 2 (1, 3) | 2 (1,3) | 0.600 |
| Antiarrhythmic Drug, n | 61 (93.8%) | 61 (93.8%) | 1.000<sup>*</sup> |
| Beta Blocker, n | 44 (67.7%) | 48 (73.8%) | 0.441 |
| Nondihydropyridine Calcium Channel Blocker, n | 4 (6.2%) | 4 (6.2%) | 0.604 |
| Hematocrit, % | 42.5 ± 4.9 | 43.8 ± 4.7 | 0.117 |
| Serum Platelet, 10<sup>9</sup>/L | 216.0 ± 55.9 | 213.0 ± 50.4 | 0.751 |
| Serum Creatinine, mg/dL | 0.9 ± 0.2 | 1.0 ± 0.8 | 0.457 |
| N-terminal Prohormone of Brain Natriuretic Peptide, pg/mL | 187.0 [51.9, 522.5] | 281.0 [78.5, 582.5] | 0.378** |
| Anteroposterior Dimension of Left Atrium on Transthoracic Echocardiography, mm | 42.6 ± 6.7 | 43.0 ± 5.7 | 0.713 |
| Left Ventricular Ejection Fraction on Transthoracic Echocardiography, % | 63.5 ± 9.3 | 63.9 ± 9.3 | 0.792 |
| Left Atrial Volume assessed using CT, ml | 107.6 [72.1, 142.2] | 107.0 [87.0, 137.2] | 0.945** |

Values are presented as number (%), mean ± standard deviation, or median [interquartile range].
\* by Fisher's exact test
\*\* by Mann–Whitney U test

### Procedural characteristics and acute outcomes

Preprocedural left atrial volume and the number of mapping points used for three-dimensional reconstruction were similar between the two groups. Cardiac rhythm during voltage mapping, the incidence of non-pulmonary vein triggers, the proportion of the left atrial surface with bipolar voltage <0.5 mV, and the frequency of cavotricuspid isthmus ablation were also comparable (Table 2).

**Table 2.**
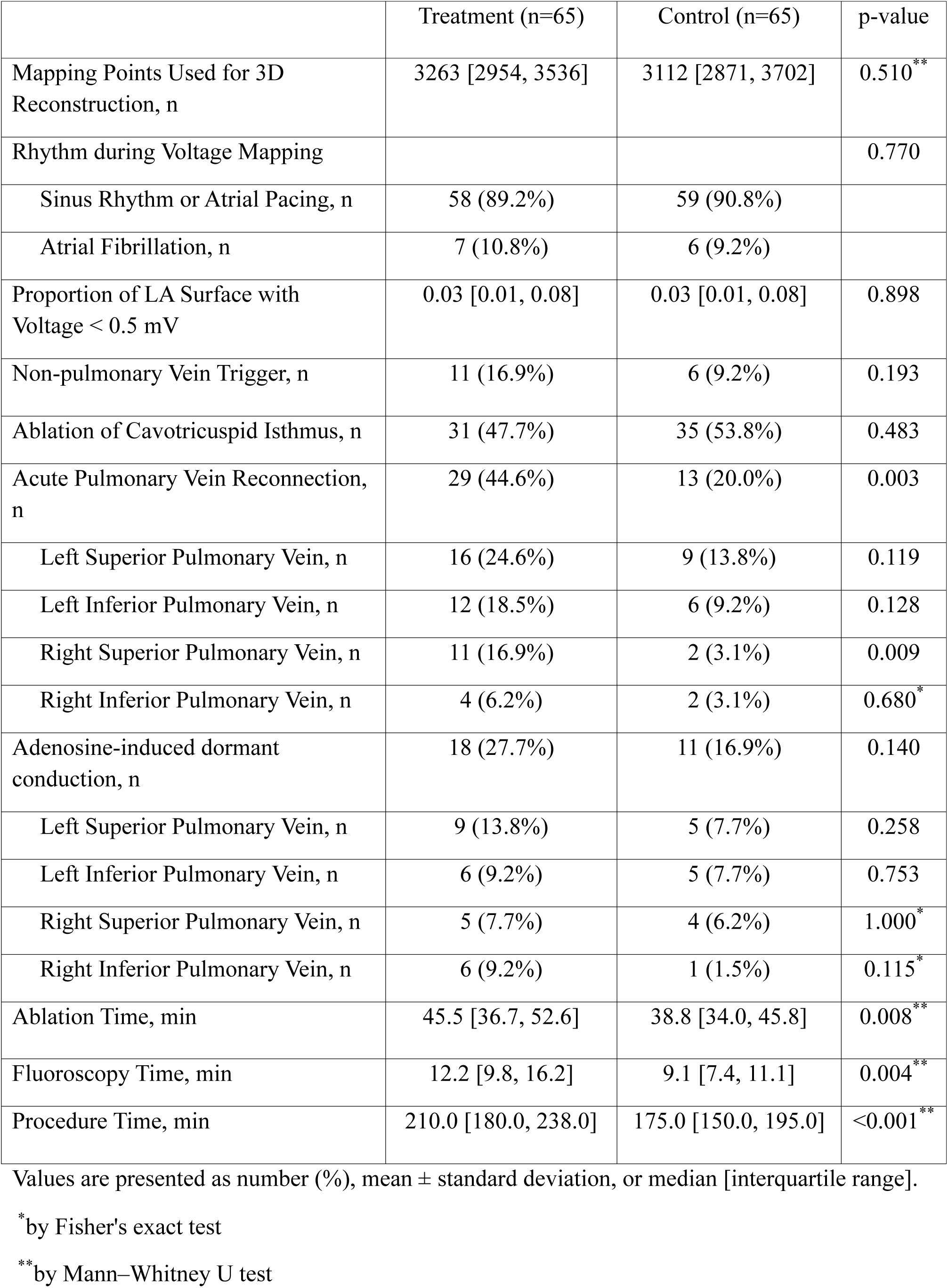
Procedure-related characteristics.

Despite these comparable anatomical and substrate characteristics, the voltage-guided stepwise pulmonary vein isolation strategy was associated with significantly longer ablation, fluoroscopy, and overall procedure times than the conventional circumferential antral pulmonary vein isolation (Table 2).

Acute pulmonary vein reconnection after completion of the initial lesion set occurred more frequently in the treatment group than in the control group (44.6% versus 20.0%; p=0.003), and this difference was most pronounced in the right superior pulmonary vein (16.9% versus 3.1%; p=0.009). The incidence of adenosine-induced dormant conduction was similar between the two groups.

### Primary outcome: 1-year atrial tachyarrhythmia recurrence

During 1 year of follow-up, atrial tachyarrhythmia recurred in 23 (35.4%) patients in the treatment group and in 13 (20.0%) patients in the control group. The absolute risk difference between groups in the 1-year recurrence rate was 15.4 percentage points (90% confidence interval 2.7–28.1). In the non-inferiority analysis, the treatment strategy was not shown to be non-inferior to the control strategy (one-sided p for non-inferiority = 0.520). In the Kaplan-Meier analysis of time to first recurrence (Figure 2), freedom from atrial tachyarrhythmia was significantly lower in the treatment group than in the control group (log-rank p = 0.033). In a Cox proportional hazards model, assignment to the treatment group was associated with a higher risk of recurrent atrial tachyarrhythmia compared with the control group (hazard ratio 2.05, 95% confidence interval 1.04–4.06; p = 0.038). A complementary non-inferiority analysis based on the hazard ratio likewise failed to demonstrate non-inferiority of the treatment strategy (one-sided p for non-inferiority = 0.429). During the total follow-up period beyond one year, this divergence between Kaplan–Meier curves persisted (Supplementary Figure 1).

**Figure 2.**
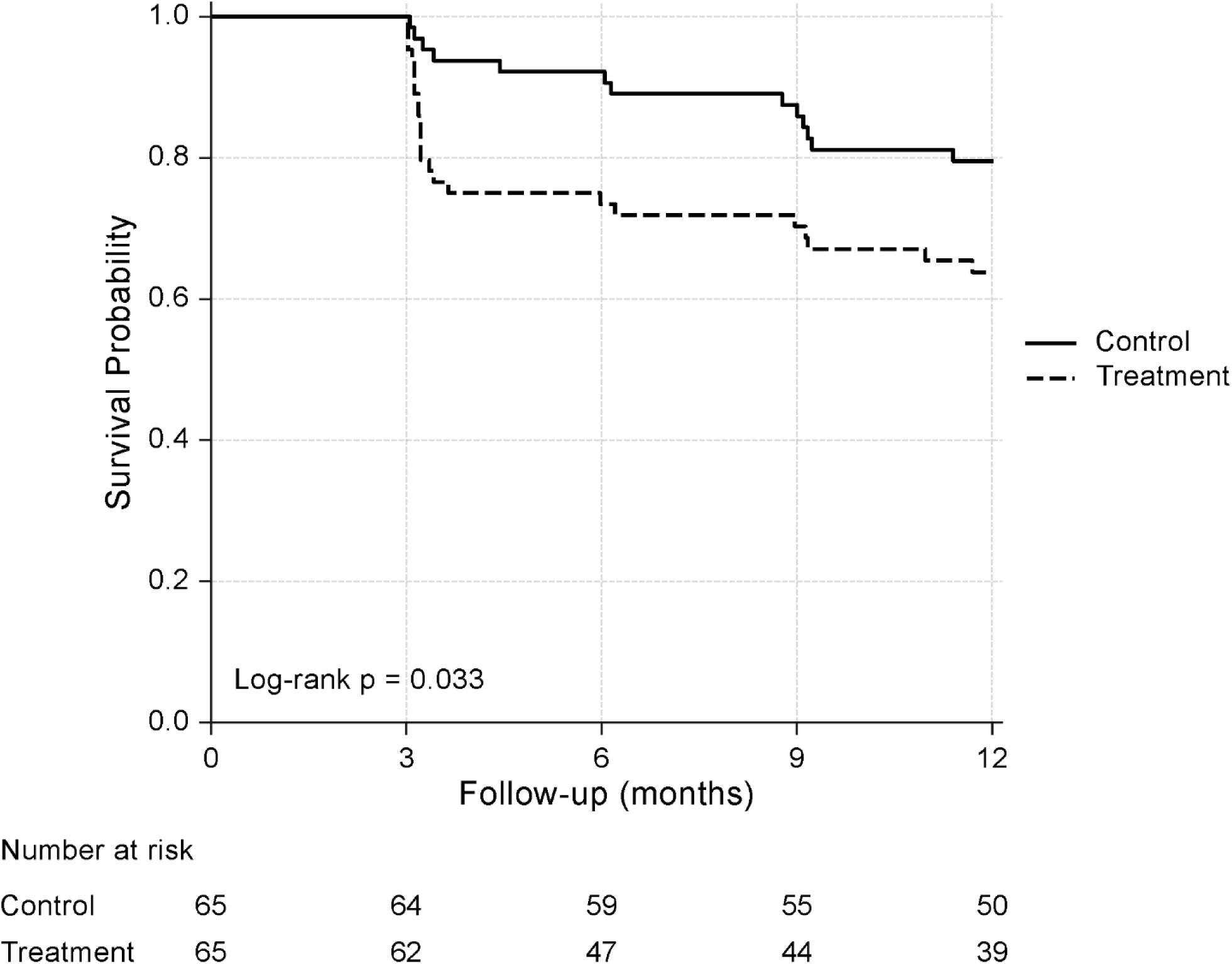
Kaplan–Meier Curve for Atrial Tachyarrhythmia Recurrence During 1-Year Follow-up

### Safety outcomes

Acute procedural complications occurred in 1 patient (1.5%) in the treatment group and in 6 patients (9.2%) in the control group (p = 0.115, Fisher’s exact test; Table 3). The only complication in the treatment group was a systemic embolism presenting as retinal artery occlusion, whereas complications in the control group included 3 cases of cardiac tamponade and 3 cases of esophageal thermal injury documented on esophagoscopy. All esophageal thermal injuries occurred in the control group without clinically overt sequelae (Supplementary Figure 2). There were no procedure-related deaths, ischemic strokes, pacemaker implantations, phrenic nerve palsy, or clinically significant hematomas at the vascular access site.

**Table 3.** Acute procedural complications.

| Complication | Treatment (n=65) | Control (n=65) | p-value |
| --- | --- | --- | --- |
| Any acute procedural complication | 1 (1.5%) | 6 (9.2%) | 0.115<sup>*</sup> |
| Death, n | 0 (0.0%) | 0 (0.0%) |  |
| Cardiac Tamponade, n | 0 (0.0%) | 3 (4.6%) |  |
| Implantation of Pacemaker, n | 0 (0.0%) | 0 (0.0%) |  |
| Palsy of Phrenic Nerve, n | 0 (0.0%) | 0 (0.0%) |  |
| Hematoma at Vascular Access Site, n | 0 (0.0%) | 0 (0.0%) |  |
| Ischemic Stroke, n | 0 (0.0%) | 0 (0.0%) |  |
| Systemic Embolism, n | 1 (1.5%) | 0 (0.0%) |  |
| Esophageal Thermal Injury, n | 0 (0.0%) | 3 (4.6%) |  |

Values are presented as number (%).
<sup>\*</sup>by Fisher's exact test

### Bipolar Voltage and Isolation Threshold

As an exploratory assessment of the voltage mapping used in this study, we first examined the relationship between bipolar and unipolar electrograms that were recorded simultaneously at identical mapping points throughout the left atrium. Bland–Altman plots and Spearman correlation coefficients showed small bias and moderate to strong correlations across all segments, and this relationship between bipolar and unipolar voltages did not differ between the treatment and control groups (Supplementary Figure 3).

Subsequently, the maximum bipolar voltage within each ablation segment was related to the bipolar voltage threshold at which pulmonary vein isolation was achieved, and a clear positive linear association was observed between these measures (Supplementary Figure 4).

In mixed-effects analyses, the estimated mean isolation voltage threshold was higher at left pulmonary veins than at right pulmonary veins (p < 0.001; Supplementary Figure 5A) and was higher in patients with paroxysmal than in those with non-paroxysmal atrial fibrillation (p = 0.012; Supplementary Figure 5B). In contrast, sex, age group, body weight, stroke risk score, renal function, N-terminal prohormone of brain natriuretic peptide category, left atrial diameter, rhythm during voltage mapping, and the proportion of low-voltage area on the left atrial surface were not consistently associated with the estimated mean isolation voltage threshold. Subject-specific random effects demonstrated wide dispersion in baseline isolation voltage threshold (Supplementary Figure 6), indicating substantial interindividual variability. These findings suggest that, at the group level, isolation voltage threshold increases with local bipolar voltage and differs according to pulmonary vein and atrial fibrillation phenotype. However, for individual patients, the exact isolation threshold cannot be reliably predicted from baseline clinical and mapping characteristics alone.

## Discussion

In this prospective, multicenter randomized trial, a voltage-guided stepwise pulmonary vein isolation strategy did not achieve non-inferiority compared with conventional circumferential antral pulmonary vein isolation in 1-year atrial tachyarrhythmia recurrence. Moreover, time-to-event analyses favored the conventional approach, with higher recurrence risk in the voltage-guided group. These results suggest that escalating lesion delivery according to local bipolar voltage does not ensure a contiguous, durable antral lesion set.

Several procedural observations provide plausible explanations for the higher recurrence rate in the voltage-guided group. Despite comparable baseline substrate characteristics and mapping conditions, the voltage-guided strategy was associated with longer procedure times. Notably, acute pulmonary vein reconnection after the initial lesion set occurred more frequently with the stepwise approach, most prominently in the right superior pulmonary vein. Recurrences clustered early during follow-up, particularly in the voltage-guided group. Because acute reconnection reflects incomplete lesion continuity or insufficient transmurality,^10,11^ a higher rate of early reconnection offers a mechanistic link to the observed excess recurrence. The stepwise design inherently emphasizes incremental lesion delivery and does not consistently yield a fully contiguous lesion set. This can leave vulnerable gaps in durable conduction block or produce nonuniform lesion density, particularly in anatomically challenging regions where catheter stability and tissue contact vary.^5,6^ In this context, a longer procedural duration may represent not only increased complexity but also a greater opportunity for tissue edema and shifting lesion geometry, which could further undermine the stability of acute pulmonary vein isolation.^12^

The exploratory voltage analyses provide additional context for interpreting these results. Agreement between bipolar and unipolar voltages was similar across treatment assignment. This finding is consistent with prior reports comparing bipolar and unipolar voltages at identical mapping sites and supports consistent voltage acquisition and annotation between groups.^13^ In addition, maximal segmental bipolar voltage showed a positive association with the isolation voltage threshold, indicating that electrogram amplitude reflects local conduction properties relevant to achieving electrical isolation. However, the mixed-effects model suggested substantial interindividual variability in the estimated isolation voltage threshold, which may limit the ability to derive individualized isolation thresholds from baseline clinical or mapping characteristics alone. Overall, these exploratory results are hypothesis generating and suggest that voltage metrics warrant evaluation alongside additional factors that influence durable pulmonary vein isolation.

Esophageal thermal injury was observed more frequently in the control group than in the voltage-guided group. Prior systematic endoscopic surveillance studies have shown that esophageal thermal injuries detected after thermal ablation are not uncommon and are frequently clinically silent.^14^ Accordingly, the observed group difference in esophageal thermal injury is consistent with the procedural premise of the voltage-guided stepwise strategy, which is expected to reduce posterior wall lesion burden relative to a fully contiguous conventional approach.

These results do not support adopting the voltage-guided stepwise strategy as an alternative approach for pulmonary vein isolation. Although voltage mapping can characterize local electrogram substrate and help interpret variability in isolation thresholds, its use should not compromise key determinants of pulmonary vein isolation durability.^15,16^ To optimally apply voltage mapping as an adjunct, future strategies may be better served by guiding targeted additional lesion delivery at regions prone to reconnection, such as the carinae, the left atrial appendage ridge, and anatomic variants including a common pulmonary vein.^17,18^ Integrating voltage with complementary intraprocedural markers, such as contact force, impedance drop, and measures of lesion contiguity, may be required to enable personalization without compromising durability.

This study has several limitations. First, operators were not blinded to group assignment, which could have influenced procedural decision making. However, randomization, use of the same predefined pre-ablation line in both groups, and standardized energy settings and procedural end points were intended to mitigate operator-related bias. Operator blinding is often challenging in procedure-based randomized trials, and performance bias cannot be fully excluded.^19^ Second, antiarrhythmic drug use during follow-up was permitted at physician discretion and could have affected recurrence detection. Nevertheless, antiarrhythmic drug use during extended follow-up was similar between groups (47.7% in the treatment group vs 40.0% in the control group, p = 0.377), making differential medication exposure an unlikely explanation for the observed outcome differences. Third, the trial was designed for a non-inferiority comparison with a prespecified margin and a target power of 70%. Even under this design, non-inferiority was not demonstrated, supporting the robustness of the primary conclusion. Finally, the voltage-related analyses were exploratory and hypothesis generating, and multiple testing and residual confounding limit causal interpretation.

In conclusion, the voltage-guided stepwise strategy did not demonstrate non-inferiority and was associated with longer procedures and more acute reconnection. Isolation voltage threshold was related to bipolar voltage and varied by anatomic location and atrial fibrillation phenotype, but interindividual variability limited individualized prediction.

## Data Availability

All data referred to in the manuscript are available within the article and its supplementary materials.

## Acknowledgments

We thank Kyoungwon Park and Do Hyeong Kim (Abbott field clinical engineers) for technical assistance during the ablation procedures.

## Disclosure

The authors report no conflicts of interest.

## Funding

This study was supported by an investigator sponsored research grant from Abbott (Investigator Sponsored Studies program; Study ID 27910 and 20898)

## Author contributions

D.-H.K., Y.-S.B., D.Y.K., G.-S.H., D.-I.L., and K.-N.L. contributed to data acquisition and data curation. D.-H.K. and Y.-S.B. drafted the manuscript as co–first authors. K.-N.L. performed the statistical analysis and critically revised the manuscript, including final review. D.-I.L. critically revised the manuscript and provided final review. All authors reviewed and approved the final manuscript.

---

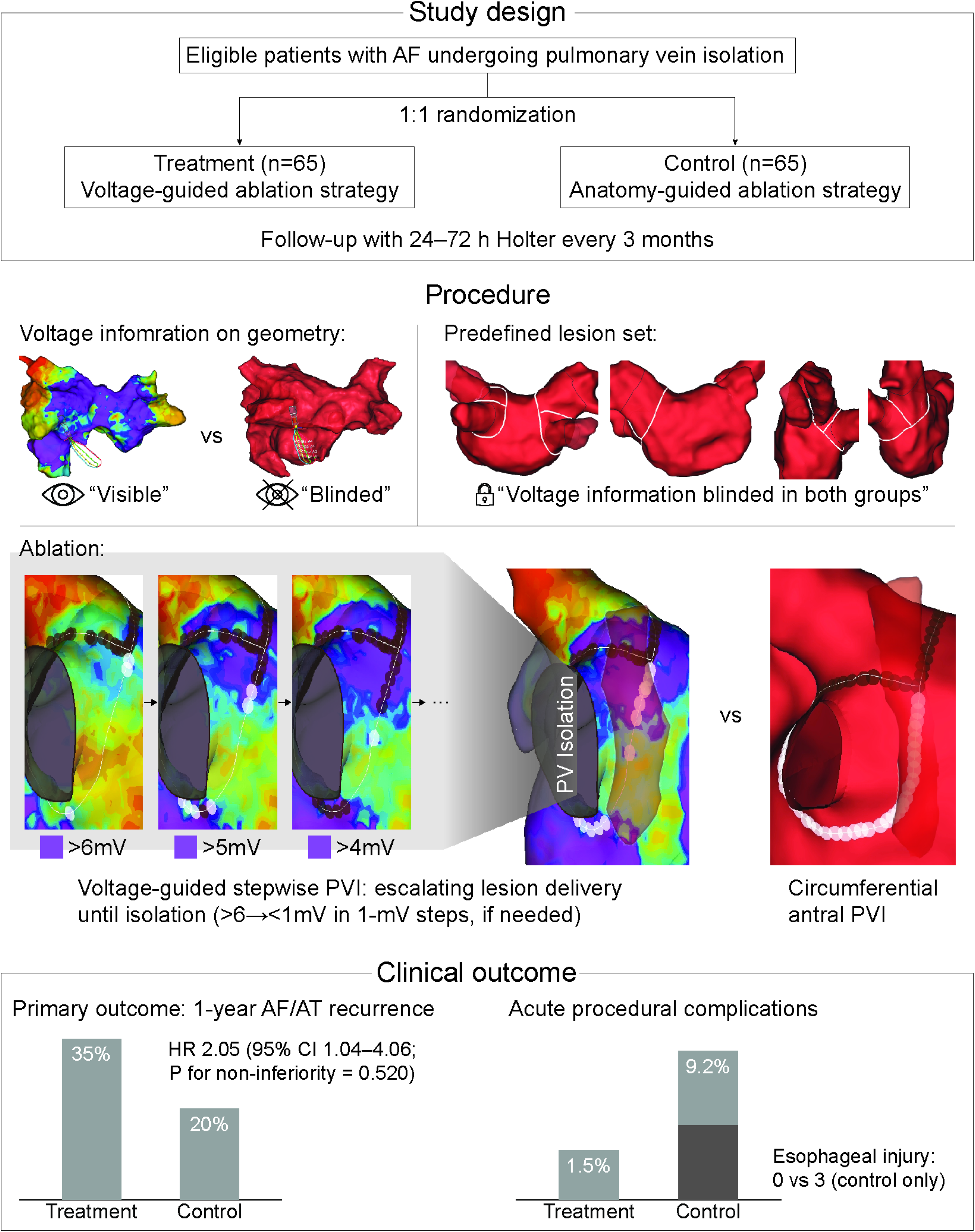

## Notes

### Competing Interest Statement

The authors have declared no competing interest.

### Clinical Trial

NCT07354737

### Author Declarations

The protocol was approved by the Institutional Review Board/ethics committee at each participating center (lead site: IRB of Ajou University Hospital approval No. AJIRB-MED-INT-20-475). Written informed consent was obtained from all participants.

